# Urban-rural differences in physical therapy use among US adults with arthritis

**DOI:** 10.64898/2025.12.16.25342395

**Authors:** Kevin H. McLaughlin, Allyn M. Bove, Kate I. Minick, Richard L. Skolasky

## Abstract

**Purpose:** The purpose of this study was to analyze differences in physical therapy (PT) use among a nationally representative sample of adults with symptomatic arthritis according to the rurality of their residence.

**Methods:** We used data from the 2023 National Health Information Survey (adult sample) to identify individuals with symptomatic arthritis by using 2 survey items indicating whether a healthcare provider had diagnosed the participant with arthritis and whether the participant had experienced arthritis-related symptoms during the past 30 days. One survey item was used to identify whether the participant had participated in PT during the previous 12 months. Levels of rurality were defined, in ascending order, as “large central metropolitan,” “large fringe metropolitan,” “medium or small metropolitan,” and “nonmetropolitan/rural,” according to each participant’s county of residence. Univariate and multivariate statistics were used to determine the association of PT use with the level of rurality. National estimates were calculated using weighting variables.

**Findings:** We identified 5,749 adults (weighted = 40,358,683) meeting our definition of symptomatic arthritis. Compared to those living in large central metropolitan areas, participants living in medium or small metropolitan areas were 20% less likely to report PT use (weighted odds ratio: 0.80; 95% confidence interval: 0.66, 0.96) and those living in nonmetropolitan/rural areas were 30% less likely (weighted odds ratio: 0.70; 95% confidence interval: 0.56, 0.88).

**Conclusions:** Among adults with symptomatic arthritis, those living in more rural areas had lower odds of PT use than those living in less rural (more urban) areas.

## INTRODUCTION

Arthritis is among the leading causes of disability in the United States, with approximately one quarter of adults older than 20 years reporting arthritis-related symptoms each year.^1^ Osteoarthritis, the most common form of arthritis, accounted for $80 billion in US healthcare spending in 2018, equal to approximately $108 billion in 2025 when accounting for inflation.^2^ Public health surveillance data suggest that the burden of arthritis is greatest among those living in rural parts of the US, 32% of whom report having been diagnosed with arthritis compared with 20% living in urban areas.^3^

Physical therapy (PT) is recommended as a first-line treatment for patients experiencing arthritis, with a large body of evidence supporting the clinical effectiveness of PT interventions (e.g., exercise, manual therapy) for reducing pain and disability.^4–8^ Access to PT may be limited in rural areas, with studies suggesting that the number of physical therapists per capital is up to 40% less than in urban communities.^9,10^ One study also found that individuals with low back pain are 34% less likely to use PT when living in rural communities.^11^ However, it is unclear whether similar urban-rural differences exist in PT use among adults with arthritis. Therefore, the purpose of this study was to analyze differences in PT use among a nationally representative sample of adults with symptomatic arthritis according to the rurality of their residence.

## MATERIALS AND METHODS

We used data from the 2023 National Health Interview Survey (NHIS) adult sample to investigate PT use among adults diagnosed with arthritis. The NHIS is an annual survey conducted by the National Center for Health Statistics, part of the US Centers for Disease Control and Prevention. It is a cross-sectional household survey that includes people living in private residences and certain noninstitutional group settings, such as group homes and homeless shelters.^12^ It does not include individuals without permanent addresses, such as active-duty military personnel and those living in correctional facilities. Interviewers conduct the NHIS in person throughout 2023, using a geographically clustered sampling approach. The sampling design ensures that the survey is nationally representative, enabling population-level estimates. Notably, the sample includes a representative number of individuals living in rural areas, which is essential for this study.

### Study Sample

We included individuals participating in the 2023 NHIS who were actively experiencing symptoms related to arthritis. We identified these individuals according to responses to 2 survey items. The first item asked participants if they had ever been told by a doctor or other healthcare professional that they had some form of arthritis. The second item asked participants if they had experienced symptoms of pain or stiffness in or around their joints (excluding the neck and back) during the previous 30 days. Participants who responded “yes” to both items were included in the study cohort. We chose this combination of items because it enabled us to identify individuals with symptomatic arthritis, who are more likely to seek care to manage their symptoms compared to individuals who have been diagnosed with arthritis but are asymptomatic.

### Physical Therapy Use

We identified PT use according to participants’ response to a question asking if they had used PT during the previous 12 months. Participants responding “yes” to this item were classified as PT users.

### Rurality

We identified participants’ level of rurality using the 2013 National Center for Health Statistics Urban-Rural Classification Scheme, a 4-level classification system used to describe the rurality of US counties and their equivalents (e.g., parishes, boroughs).^13^ The level of rurality was categorized, in ascending order, as follows: 1) large central metropolitan, 2) large fringe metropolitan, 3) medium and small metropolitan, and 4) nonmetropolitan/rural.

### Covariates

We extracted demographic data from the NHIS dataset to account for individual-level factors that may have affected participants’ use of PT beyond rurality. These factors were patient age (categorized as 18–44, 45–64, and ≥65 years), health insurance coverage (yes/no), educational level (less than high school, high school/some college, associate’s degree, bachelor’s degree or higher), and income to poverty threshold ratio (based on family size, as included in the NHIS base dataset).

### Data Analysis

To appropriately address the NHIS’s multistage sampling framework, which incorporates both stratification and clustering, we conducted survey-weighted analyses and applied the sampling information provided in the NHIS files (including primary sampling units, strata, and survey weights). We used descriptive statistics to summarize characteristics of the study population. To assess the relationship between rural residence (independent variable) and PT use (dependent variable), we performed logistic regression analyses that included all covariates described above. Odds ratios were deemed significant when their 95% confidence intervals excluded 1. Consistent with National Center for Health Statistics recommendations, NHIS survey weights were applied to generate nationally representative estimates.^12^ All analyses were conducted using Stata statistical software, release 17 (StataCorp LLC; College Station, TX).

## RESULTS

We identified 5,749 individuals (weighted = 40,358,683) who met our eligibility criteria for symptomatic arthritis (Table 1). Included individuals were most commonly 65 years of age or older (59.8% of sample, 50.2% weighted) and female (62.2% of sample, 60% weighted). Participants most commonly identified as white (82.1% of sample, 80.5% weighted) and non-Hispanic (92.2% of sample, 90.8% weighted). The most common urban-rural level was medium or small metropolitan (33.8% of sample, 32.7% weighted) and the least common was nonmetropolitan/rural (20.7% of sample, 19.1% weighted).

**Table 1.** Characteristics of US adults reporting symptomatic arthritis, 2023 National Health Interview Survey.

| Characteristic | N (%) |  |
| --- | --- | --- |
|  | Sample (n = 5,749) | Weighted (n = 40,358,683) |
| Age category, yr |  |  |
| 18–44 | 457 (8.0) | 4,587,933 (11.4) |
| 45–64 | 1,850 (32.2) | 15,496,918 (38.4) |
| ≥65 | 3,437 (59.8) | 20,255,414 (50.2) |
| Female sex | 3,576 (62.2) | 24,022,129 (60.0) |
| Race |  |  |
| Asian | 126 (2.2) | 1,173,764 (2.9) |
| Black | 595 (10.4) | 4,229,112 (10.5) |
| Native American or Alaskan Native | 42 (0.7) | 271,678 (0.7) |
| White | 4,718 (82.1) | 32,497,340 (80.5) |
| Other race/multiracial | 114 (2.0) | 788,320 (2.0) |
| Unknown | 154 (2.) | 1,398,479 (3.5) |
| Ethnicity |  |  |
| Hispanic | 445 (7.7) | 3,681,353 (9.1) |
| Non-Hispanic | 5,299 (92.2) | 36,637,680 (90.8) |
| Unknown | 5 (0.1) | 39,651 (0.1) |
| Urban-rural level |  |  |
| Large central metropolitan | 1,362 (23.7) | 9,752,603 (24.2) |
| Large fringe metropolitan | 1,256 (21.9) | 9,689,567 (24.0) |
| Medium or small metropolitan | 1,943 (33.8) | 13,207,138 (32.7) |
| Nonmetropolitan/rural | 1,188 (20.7) | 7,709,376 (19.1) |

We observed that 28.8% (sample n=1,652) of our sample and 27.6% of our weighted sample (weighted n=11,137,012) reported participating in PT during the previous 12 months (Table 2). The proportion of individuals reporting PT use was the highest among those living in large central metropolitan areas (32.4% of sample, 31.3% weighted). The proportion of individuals attending PT declined with increasing levels of rurality, with the lowest rate of PT use observed among those living in nonmetropolitan/rural areas (25.2% of sample, 23.4% weighted).

**Table 2.**
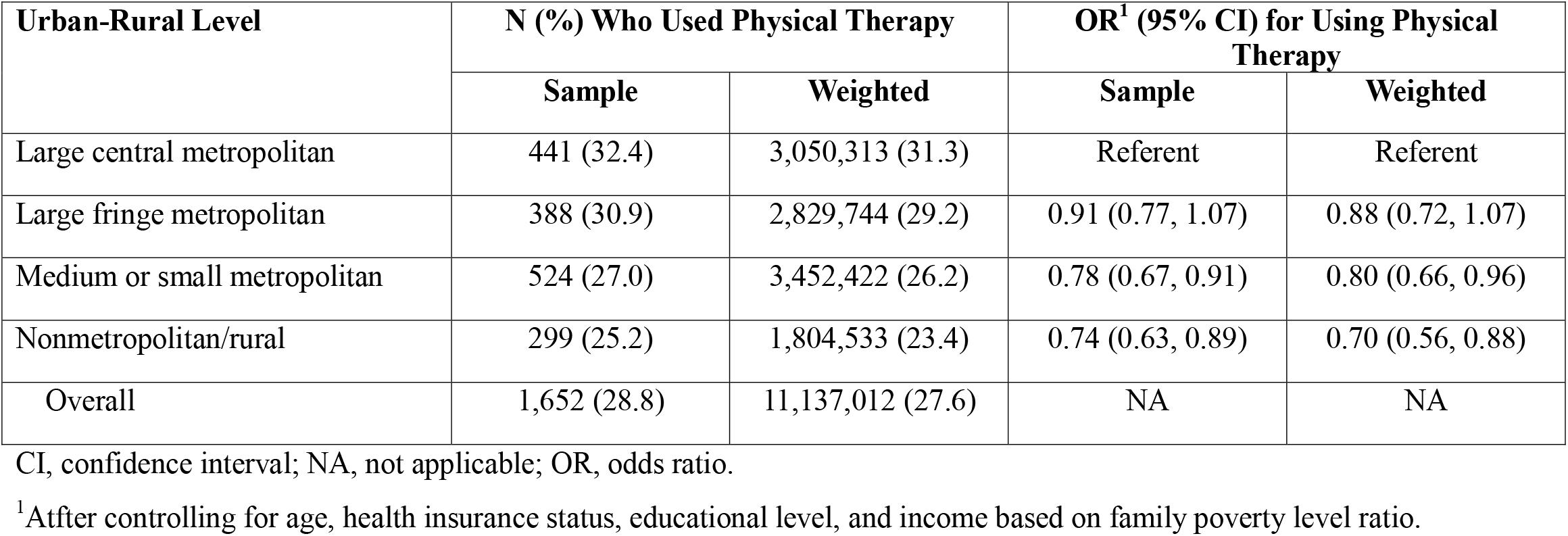
Self-reported physical therapy use among adults with symptomatic arthritis, 2023 National Health Interview Survey.

| Urban-Rural Level | N (%) Who Used Physical Therapy |  | OR<sup>1</sup> (95% CI) for Using Physical Therapy |  |
| --- | --- | --- | --- | --- |
|  | Sample | Weighted | Sample | Weighted |
| Large central metropolitan | 441 (32.4) | 3,050,313 (31.3) | Referent | Referent |
| Large fringe metropolitan | 388 (30.9) | 2,829,744 (29.2) | 0.91 (0.77, 1.07) | 0.88 (0.72, 1.07) |
| Medium or small metropolitan | 524 (27.0) | 3,452,422 (26.2) | 0.78 (0.67, 0.91) | 0.80 (0.66, 0.96) |
| Nonmetropolitan/rural | 299 (25.2) | 1,804,533 (23.4) | 0.74 (0.63, 0.89) | 0.70 (0.56, 0.88) |
| Overall | 1,652 (28.8) | 11,137,012 (27.6) | NA | NA |

CI, confidence interval; NA, not applicable; OR, odds ratio.
<sup>1</sup> After controlling for age, health insurance status, educational level, and income based on family poverty level ratio.

Based on multivariable analysis, individuals living in medium and small metropolitan areas had 20% lower odds (weighted odds ratio: 0.80; 95% confidence interval: 0.66, 0.96) of reporting PT use over the past 12 months compared to similar individuals living in large central metropolitan areas. Individuals living in nonmetropolitan/rural areas had 30% lower odds (odds ratio: 0.70; 95% confidence interval: 0.56, 0.88) of reporting PT use during this timeframe compared to those living in central metropolitan areas.

## DISCUSSION

This study is one of the first to examine the association of rurality with PT use among individuals with arthritis. The results indicate that greater levels of rurality are associated with lower odds of PT use among adults with symptomatic arthritis. At the far end of the spectrum, we observed that individuals living in nonmetropolitan/rural parts of the US had 30% lower odds of reporting PT use during the past 12 months compared to those living in large central metropolitan areas. These findings represent potential urban-rural disparities in access to PT among US adults with arthritis and warrant further investigation into the factors that affect PT use among urban and rural populations.

The results of this study are consistent with those of a previous study investigating the influence of rurality on PT use among individuals with severe chronic low back pain using data from the 2019 NHIS.^11^ That study found that individuals with severe chronic low back pain living in nonmetropolitan/rural areas were 34% less likely to report using PT during the previous 3 months compared to individuals living in large central metropolitan areas. It is possible that lower rates of PT use among rural individuals with low back pain are related to a preference for chiropractic care among this population.^14^ However, chiropractic care is less commonly used for joint pain outside the neck and back regions and, as such, is less likely to have influenced rates of PT use among the current study’s population, comprising individuals with extremity joint pain. Taken together, the results of these studies indicate that systematic factors likely influence the use of PT in rural areas.

Several factors could influence PT use in rural parts of the country, and research is needed to inform strategies for improving access to PT in rural communities. For example, it is likely that the lower number of physical therapists available per capita in rural parts of the US contributes to lower rates of PT use^9^ if patients must travel farther to receive PT or if distance makes it unfeasible for them to attend PT at all. PT conducted by telehealth is viewed by many physical therapists and patients as an acceptable alternative to in-clinic PT and may represent a strategy for overcoming geographical barriers to PT care.^15,16^ However, telehealth is less likely to address barriers related to mistrust of healthcare providers or perceived need for healthcare services, both of which have been found to differ by rurality.^17–19^ Strategies for decreasing urban-rural healthcare disparities among those with arthritis should be based on the specific factors identified and developed in partnership with individuals living in rural communities.

Strengths of this study include use of a large, nationally representative dataset and rigorous survey methodology. One limitation to note is that we were unable to differentiate between types of arthritis diagnosed in our sample. However, PT is commonly used by individuals with several types of arthritis and is recommended as a first-line treatment for many rheumatic conditions, so we consider it unlikely that arthritis type varied greatly by rurality or had a meaningful influence on the results.

## CONCLUSION

Individuals with symptomatic arthritis who live in rural areas of the US are significantly less likely to use PT compared to those living in large central metropolitan areas. The results of this study suggest potential disparities in access to PT among individuals living in rural communities. However, further research is needed to examine the factors that influence PT use in rural areas and to develop strategies that address barriers to PT use in these communities.

## Data Availability

All data produced are available online at https://www.cdc.gov/nchs/nhis/documentation/2023-nhis.html

## Acknowledgments

For editorial assistance, we thank Rachel Walden, MS, in the Editorial Services group of The Johns Hopkins Department of Orthopaedic Surgery.

## Notes

**Disclosures:** The authors declare no conflicts of interest.

### Competing Interest Statement

The authors have declared no competing interest.

### Funding Statement

This study did not receive any funding

### Author Declarations

This study utilized publicly available data from the 2023 National Health and Interview Survey

